# Stratified cohorts for biomarker assessment and trial readiness: *TMEM175*, *SCARB2* and *CTSB* in Parkinson’s disease

**DOI:** 10.64898/2026.06.23.26356322

**Authors:** Wenhua Sun, Isabel Wurster, Benjamin Roeben, Roswitha Kemmner, Michele Mielke, Henrik Zetterberg, Stefanie Lerche, Ann-Kathrin Hauser, Claudia Schulte, Piero Parchi, Gabor C. Petzold, Annika Spottke, Ullrich Wüllner, Christoph van Riesen, Fabian Maass, Björn Falkenburger, Bähr Mathias, Inga Zerr, Emrah Duezel, Paul Lingor, Andreas Wolff, Johannes Levin, Wiebke Hermann, Mathias Löhle, Ziv Gan-Or, Kathrin Brockmann, Thomas Gasser

## Abstract

**Background:** Lysosomal dysfunction plays a crucial role in the pathogenesis of Parkinson’s disease (PD), particularly among *GBA1* mutation carriers. Beyond *GBA1*, genes such as *TMEM175*, *SCARB2*, and *CTSB* identified in genome-wide association studies (GWAS) are also implicated in lysosomal pathways contributing to PD risk, although their functional effects in patients remain unclear. Proteins encoded by these lysosome-related genes have been explored as potential therapeutic targets in experimental models. Biomarker profiles, including clinical measures, α-syn seeding activity, lysosomal proteins, and sphingolipids, may facilitate patient stratification and support therapeutic monitoring in future clinical trials.

**Aim:** The aim of this study is to investigate the impact of genetic variants of three lysosomal-related genes (*TMEM175*, *SCARB2*, and *CTSB*) on biomarker profiles in PD with and without *GBA1* variants. Cross-sectional data from two German cohorts: the Tuebingen Parkinson Cohort (TUEPAC) and the DESCRIBE PD cohort of the German Center for Neurodegenerative Diseases were used as explorative cohorts, and data from Accelerating Medicines Partnership® Parkinson’s Disease (AMP-PD) were used as a validation cohort. The ultimate goal is to provide new data for patient stratification based on genetics, which might serve as a readout for target engagement and treatment efficiency assessment.

**Methods:** Three cohorts were analyzed: TUEPAC, DESCRIBE PD, and AMP-PD. TUEPAC and DESCRIBE PD were combined into a single German discovery cohort (TUEPAC-DESCRIBE-PD), while AMP-PD served as an independent validation cohort. Within each cohort, for subgroup analyses, PD patients were classified as the overall PD cohort (PD_all_), and further stratified by *GBA1* mutation status into PD patients without *GBA1* mutations (PD_*GBA1*_wildtype_), and PD patients carrying *GBA1* mutations (PD_*GBA1*_). We evaluated cognitive and motor function, as well as depression using the Montreal Cognitive Assessment (MoCA), Unified Parkinson Disease Rating Scale-part I II (UPDRS III), and Beck Depression Inventory-II(BDI-II) scales. Analyzed biomarkers included CSF α-syn seeding activity using seed amplification assay (SAA), CSF lysosomal protein levels of lysosomal integral membrane protein 2 (LIMP2), also known as SCARB2, cathepsin B (CTSB) and lysosome-associated membrane protein 2 (LAMP2), blood-based enzyme activity of the lysosomal glucocerebrosidase (GCase), and CSF sphingolipid profiles. PD patients carrying risk alleles in *TMEM175*, *SCARB2*, and *CTSB* were compared to non-carriers.

**Results:** Genotype-phenotype correlation analysis in TUEPAC-DESCRIBE-PD and AMP-PD revealed: (1) In PD_all_, the *TMEM175* p.M393T risk variant was nominally associated with decreased cognitive function when adjusted for *GBA1* mutation status in TUEPAC-DESCRIBE-PD; this association could not be replicated, although a similar trend was observed in the slightly smaller, but multicentric AMP-PD cohort; *TMEM175* p.M393T was not significantly associated with BDI-I I or UPDRS-III scores in either cohort. (2) In PD_*GBA1*_wildtype_, GCase activity was significantly lower in PD patients with *SCARB2* rs6812193 risk allele in TUEPAC-DESCRIBE-PD, while a similar but non-significant trend was observed in AMP-PD; (3) In PD_all_, CSF levels of CTSB were nominally lower in carriers of CTSB rs1293298 risk allele compared to carriers of *CTSB* rs1293298 protective allele in TUEPAC-DESCRIBE-PD; in PD_*GBA1*_wildtype_, LAMP2 was significantly lower in carriers of *CTSB* rs1293298 risk allele compared to carriers of *CTSB* rs1293298 protective allele in TUEPAC-DESCRIBE-PD; (4) In PD_all_, *TMEM175* p.M393T risk allele was nominally associated with altered sphingolipid profiles across both TUEPAC-DESCRIBE-PD and AMP-PD cohorts.

**Conclusion:** These findings demonstrate that genetic variants in lysosomal-related genes (*TMEM175*, *SCARB2*, and *CTSB*) have a functional impact on biomarker profiles in PD patients. Integrating genetic characterization with biochemical profiling provides a framework for patient stratification and may serve as a translational strategy to monitor target engagement and evaluate treatment efficacy in future clinical trials.

## Introduction

Heterozygous variants in the glucocerebrosidase gene (*GBA1*), which encodes the lysosomal enzyme β-glucocerebrosidase (GCase), represent the most common genetic risk factor for Parkinson’s disease (PD) associated with lysosomal dysfunction^1^. Reduced GCase activity disrupts sphingolipid metabolism and promotes accumulation of alpha-synuclein(α-syn)^2, 3^. Beyond the well-established role of *GBA1*, large scale genome-wide association studies (GWAS) of PD in the European population identified other genetic risk variants across lysosomal related genes: transmembrane protein 175 [ *TMEM175*, p.M393T (rs34311866) and p.Q65P (rs34884217)], scavenger receptor class B member 2 (*SCARB2*, rs6812193 and rs6825004), and cathepsin B(*CTSB*, rs1293298)^4^.

TMEM175 is a proton-selective ion channel on the lysosomal membrane, and it mediates proton efflux, accounting for over 90% of lysosomal proton flow, and thereby plays a critical role in maintaining lysosomal pH homeostasis^5^. *TMEM175* deficiency disrupts lysosomal pH and impairs TMEM175 localization, resulting in decreased enzyme activity of cathepsin D (CTSD), CTSB and GCase, and impaired autophagosome clearance, ultimately facilitating α-syn aggregation in cell models^6, 7^. In mice, deficiency in *TMEM175* leads to loss of dopaminergic neurons and motor impairment^8^. Both variants (2025 PD GWAS, p.M393T, OR =1.19; p.Q65P, OR = 0.89) have been previously reported to be associated with risk, age of onset, progression and GCase activity in PD^9–14^.

Lysosomal integral membrane protein 2 (LIMP2), encoded by *SCARB2*, transports GCase from the endoplasmic reticulum to the lysosome. LIMP2 deficiency reduced lysosomal GCase activity, accompanied by lipid accumulation and autophagic dysfunction, and α-syn accumulation in mice^15^. Two common intronic variants, rs6812193 and rs6825004 (2025 PD GWAS, rs6825004, OR = 0.96; rs6812193, OR = 0.93), were significantly associated with PD risk in several genetic studies^4, 14, 16–18^.

Cathepsin B, encoded by *CTSB*, is a lysosomal hydrolase of the cysteine cathepsin family that is normally localized to the lysosomal lumen^19^. CTSB loss of function leads to decreased GCase activity and broad lysosomal impairment, resulting in enhanced α-syn aggregation in cell lines and human dopaminergic neurons^20^. CTSB has been reported to mediate the clearance of fibrillar α-syn and maintain lysosomal function in both cellular models and in animals^20, 21^. A common intronic variant, rs1293298 (2025 PD GWAS, OR = 0.93), was identified as a genetic risk factor for PD^4, 14, 20^. This variant was a genetic modifier of disease penetrance in *GBA1* carriers, suggesting a potential genetic interaction between *CTSB* and *GBA1*^22^.

Given the established functional interactions of *TMEM175*, *SCARB2*, and *CTSB* with *GBA1* in lysosomal pathways implicated in PD pathogenesis, we aimed to perform an in-depth genetic characterization and to further examine clinical outcomes, α-syn seeding activity, lysosomal protein levels, and sphingolipid profiles associated with genetic variants in these three lysosomal related genes in PD patients from TUEPAC-DESCRIBE-PD and AMP-PD. Such genetic characterization is an essential prerequisite for patient stratification in clinical trials. Moreover, the biomarker profiles may help to identify biomarkers for target engagement and treatment efficacy assessment.

## Methods

Three independent cohorts were analyzed (Figure 1): the Tuebingen Parkinson Cohort (TUEPAC), the German Center for Neurodegenerative Diseases DESCRIBE-PD cohort, and the Accelerating Medicines Partnership Parkinson’s Disease (AMP-PD) cohort. For the present analyses, TUEPAC and DESCRIBE-PD were combined and were referred to throughout the manuscript as the discovery cohort (TUEPAC-DESCRIBE-PD). The AMP-PD cohort was used as an independent validation cohort.

**Figure 1.**
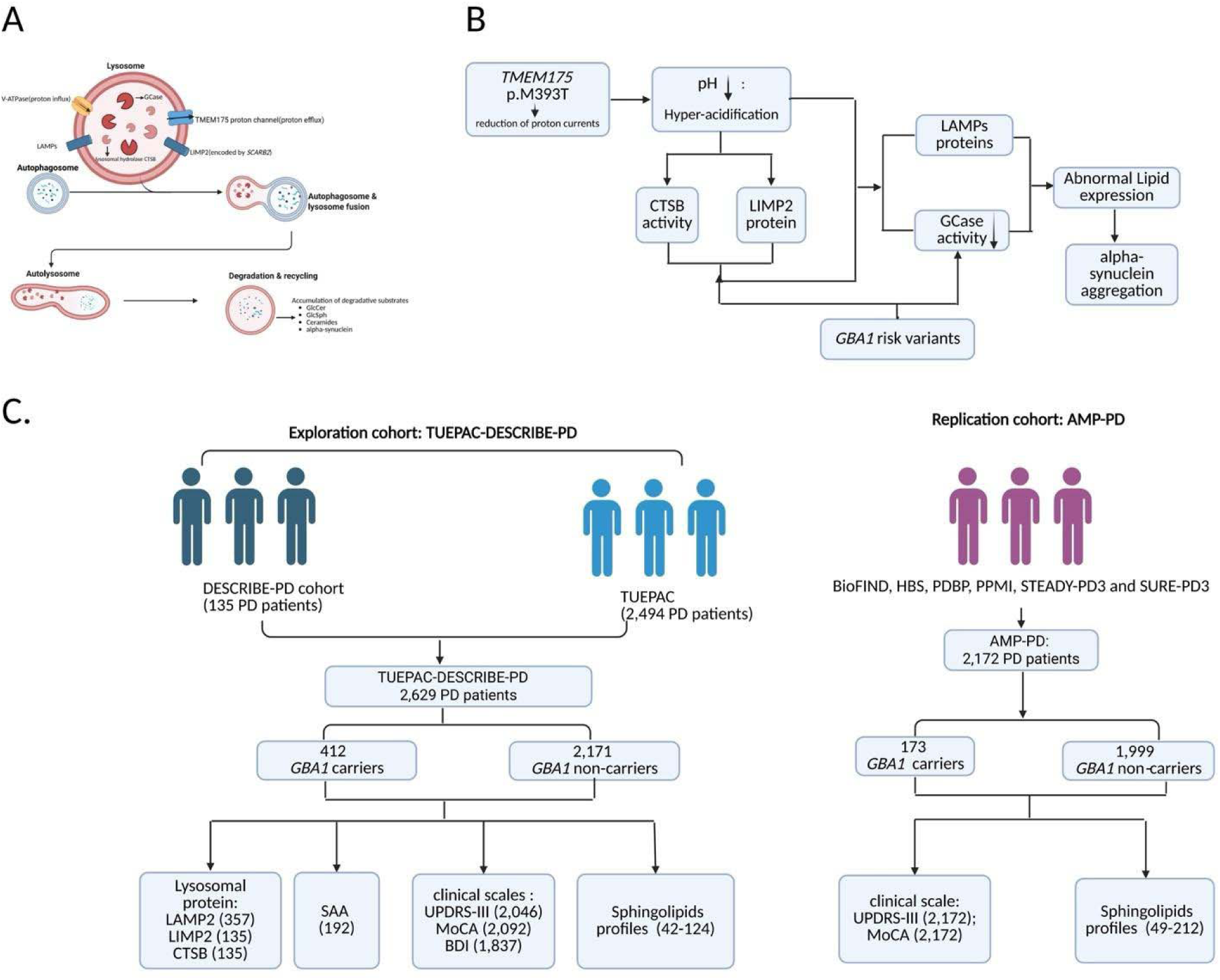
Overview of the study. A) The location of lysosomal proteins (TMEM175, SCARB2, and CTSB). B) The pathway illustrates how lysosomal proteins (TMEM175, SCARB2, and CTSB) interact with GBA1 and contribute to lysosomal dysfunction, leading to lipid accumulation and alpha-synuclein aggregation in PD. C) Cohort description of the data availability in the Tuebingen Parkinson Cohort (TUEPAC) and German Center for Neurodegenerative Diseases (DESCRIBE-PD) and Accelerating Medicines Partnership® Parkinson’s Disease (AMP-PD). Abbreviations: SAA, alpha-synuclein seed amplification assay; BioFIND, The Fox Investigation for New Discovery of Biomarkers; HBS, Harvard Biomarkers Study; PDBP, Parkinson’s Disease Biomarkers Program; PPMI, Parkinson’s Progression Markers Initiative; STEADY-PD3, Safety, Tolerability, and Efficacy Assessment of Isradipine for Parkinson’s Disease; SURE-PD3, Study of Urate Elevation in Parkinson’s Disease, Phase 3; LAMP2, Lysosome-associated membrane protein 2; LIMP2, Lysosomal integral membrane protein 2; CTSB, Cathepsin B; UPDRS-III, Unified Parkinson’s Disease Rating Scale, Part III; MoCA, Montreal Cognitive Assessment.

### 1. Discovery cohort

#### (1) Participants

2,629 PD patients were recruited from 2003 to 2024 at the University hospital of Tuebingen and other German DZNE sites (Bonn, Dresden, Göttingen and Magdeburg, München and Rostock). TUEPAC cohort was described as published previously^1^. For the new measurements, in the DESCRIBE-PD cohort the study was approved by the local ethics committee (2026-0191-BO). Patients with rare mutations in *LRRK2*, *Parkin*, *PINK1*, and *SNCA* were excluded to reduce genetic confounding effects.

#### (2) Genetic analysis

Genetic screening for *GBA1* variants was done by Sanger sequencing of the whole gene.

For 454 PD patients in the discovery cohort, genetic variants of *TMEM175*(rs34311866 and rs34884217), *SCARB2*(rs6812193 and 6825004), and *CTSB* (rs1293298) were genotyped using the SNaPshot Multiplex System (Applied Biosystems). Primers and PCR conditions are available upon request.

For an additional 2,175 PD patients in the discovery cohort, genotype data for all known genetic variants in *TMEM175*, *SCARB2* and *CTSB* genes were extracted from the GP2 Neurobooster array.

#### (3) Clinical Investigations

The UK PD Brain Bank diagnostic criteria were used for PD diagnosis^23^. Patients were assessed in dopaminergic ON. The severity of motor symptoms was evaluated using the Unified Parkinson’s disease Rating Scale-I II (UPDRS-III)^24^. Cognitive function was assessed using the Montreal Cognitive Assessment (MoCA)^25^ and the Mini–Mental State Examination (MMSE)^26^. As the MoCA was introduced only after 2009, earlier MMSE scores were converted to MoCA-equivalent scores^27^. A MoCA score of 26 or higher was considered normal cognition, whereas a score below 26 suggested cognitive impairment^28^. Depressive symptoms were assessed using the Beck Depression Inventory II (BDI-II) ^29^.

#### (4) Cerebrospinal Fluid (CSF) collection

Spinal tap was performed between 9:00 AM and 1:00 PM. Samples were directly taken from the bedside and centrifuged within 60 minutes and frozen at −80°C within 90 minutes according to previous descriptions^30, 31^.

#### (5) CSF α-syn seed amplification assay (SAA)

(A) Briefly, 15 µL of CSF was mixed with 85 µL of reaction buffer per well in a black, clear-bottom 96-well plate (Thermo Fischer Scientific), with each well containing six 0.8 mm silica beads (OPS Diagnostics). The reaction buffer consisted of 40 mM phosphate buffer (pH 8.0), 0.0015% SDS, 10µM Thioflavin T, 170 mM NaCl, and 0.1 mg/ml recombinant monomeric α-syn previously filtered through a 100 kDa MWCO centrifugal filter. Recombinant monomeric α-syn was produced as previously described^32^ at the Institute of Neurological Science of Bologna. (B) Plates were then incubated at 42°C in a FLUOstar Omega plate reader (BMG Labtech) with alternating cycles of 1 min double-orbital shaking at 400 rpm and 1 min rest. (C) Fluorescence was measured every 45 min with 448 nm excitation and 482 nm emission filter. Four technical replicates were run per sample, and each plate included two positive and two negative controls (each in quadruplicate). (D) A replicate was considered positive if its fluorescence exceeded a defined threshold within the first 30 hours. We calculated the threshold for each plate as the fluorescent value corresponding to 30% of the median fluorescence peak observed in positive replicates of the LBD control sample. (E) A sample was considered synSAA+ and assigned the “LBD-like” status if at least three replicates out of four were positive. Samples giving one or two positive replicates were repeated three times and considered synSAA+ if the yield at least 4 positive replicates out of 12. A sample was considered synSAA- and given the “non-seeder” status if none of the four replicates or less than 4 of 12 replicates (for those repeated three times) were positive.

#### (6) Measurement of lysosomal protein and sphingolipids profiles

CSF lysosomal-associated membrane protein 2 (LAMP2) protein concentration was quantified using targeted mass spectrometry (parallel reaction monitoring) with isotope-labeled internal standards, as previously described^31^.

CSF LIMP2 protein concentration was measured using human LIMP-II (SCARB2) ELISA kit (Thermo Fisher Scientific, Cat No: EHSCARB2, USA). CSF CTSB concentration was analyzed by Cathepsin B Human ELISA kit (Abcam, Cat No: ab119584, UK). All assays were performed in duplicate according to the manufacturers’ instructions.

Lysosomal GCase enzyme activity was determined in fresh blood leukocytes by assessing the hydrolytic degradation of GlcCer, as previously described^1, 33, 34^.

CSF levels of GCase upstream substrate, glucosylceramides (GlcCer), were measured on a Thermo Quantiva mass spectrometer coupled with a Waters Acquity UPLC system (Milford, MA)^1, 35^. CSF levels of GCase downstream product, ceramides (Cer), and by-products sphinganine (SPA), sphingosines (Sph), and sphingosine-1-phosphate (S1P) were measured using a Thermo TSQ Quantum Ultra mass spectrometer (West Palm Beach,FL) coupled with a Waters Acquity UPLC system (Milford, MA)^1, 36^.

### 2. The validation cohort

#### (1) Participants

The AMP-PD has developed a research platform for PD that integrates the storage and analysis of whole-genome sequencing (WGS) data, clinical data, and molecular data, harmonized across multiple cohort studies. For the present analyses, 2,172 PD patients from AMP-PD were included.

Participants’ clinical information and genetic samples were obtained under appropriate written consent and with local institutional and ethical approvals. The details of these studies can be obtained from the AMP PD website (https://amp-pd.org) and each study website.

#### (2) Genetic analysis

Information about the processing and quality checks performed on the AMP-PD WGS data can be found at https://amp-pd.org/whole-genome-data. WGS data in AMP-PD underwent sample-level and genetic quality control. Sample quality checks included assessment of contamination (Freemix < 3%), coverage (mean coverage < 25), WGS metric outliers (transition/transversion ratio < 2), and missingness (missingness genotype rates per sample > 5%). Genetic data checks included duplication check, concordance against NeuroX data, sex concordance check, excessive heterogeneity (F > +/− 0.15), and genetic ancestry outlier check using principal component analysis.

WGS data from AMP-PD participants were used to classify *GBA1* genetic variants consistent with TUEPAC-DESCRIBE-PD. Genotypes at *TMEM175* (rs34311866 and rs34884217), *SCARB2* (rs6812193), and *CTSB* (rs1293298) were extracted from the AMP-PD WGS dataset. Consistent with discovery cohort, PD patients with rare mutations (*LRRK2*, *PINK1*, *PRKN*, and *SNCA*) were excluded.

Genotype at *SCARB2* rs6825004 was not available in the AMP-PD WGS dataset.

#### (3) Clinical Investigations

PD diagnosis was based on the United Kingdom PD Brain Bank Diagnostic Criteria^23^. Motor severity was assessed using the UPDRS Part I II scores^24^. Cognitive function was assessed using the Montreal Cognitive Assessment (MoCA)^25^. The same MoCA cutoff used in TUEPAC-DESCRIBE-PD was applied to the AMP-PD dataset.

#### (4) Measurement of sphingolipids profiles

The activities of the lysosomal enzyme GCase was measured in CSF using fluorogenic substrates, according to previously published procedures^37, 38^. Like previous work^39^, plasma levels of selected sphingolipids in the *GBA1* pathway were quantified using targeted liquid chromatography-tandem mass spectrometry (LC-MS/MS) and complementary biochemical assays.

## Statistics

Based on the risk allele frequency, individuals were defined as follows: when the risk allele frequency was > 0.5, risk allele homozygous carriers were classified as risk allele carriers, while homozygous and heterozygous non-risk-allele carriers were considered protective. When the risk allele frequency was <0.5, both homozygous and heterozygous carriers of the risk allele were classified as risk allele carriers.

(1) Chi-square test was used to compare the distributions of *GBA1* carrier status (carriers vs non-carriers) and sex across all the examined genetic variants of lysosomal genes (*TMEM175*, *SCARB2* and *CTSB*).

(2) Intergroup comparisons of continuous variable (age) were done using two-sample t-test. Mann–Whitney U test was used to compare disease duration between groups.

(3) Given that *GBA1* carriers may influence clinical outcomes^40^, α-syn seeding activity^30^, lysosomal protein levels^31^ and sphingolipid profiles^1^, we adopted two ways to adjust for this possible confounder. On one hand, *GBA1* carrier status was included as a covariate. Multivariable linear regression, adjusted by sex and *GBA1* status, was used to assess associations with age at onset and disease duration between the groups. Multivariable linear regression, adjusted by sex, age at visit, disease duration and *GBA1* status, was used to assess genotype-phenotype associations for clinical outcomes (UPDRS-III and BDI-II), lysosomal proteins (LAMP2, CTSB and LIMP2) and sphingolipids profiles between the groups. Multivariable logistic regression was applied to assess associations between genetic variants and categorical outcomes (MoCA and a-syn seeding activity). On the other hand, participants were stratified into *GBA1* subgroups (PD_*GBA1*_wildtype_ and PD_*GBA1*_) to account for potential *GBA1*-related effects and regression analyses were conducted to assess differences between carriers of risk and protective alleles across lysosomal related genes (*TMEM175*, *SCARB2* and *CTSB*), adjusting for sex, age at visit and disease duration.

(4) Spearman’s rank correlation was used to evaluate associations among the study variables, including UPDRS-III, MoCA, BDI-I I, a-syn seeding activity, lysosomal protein levels and sphingolipids profiles.

(5) To account for multiple testing, raw p-values were adjusted using the Benjamini–Hochberg (BH) method to control false discovery rate (FDR) across five genetic variants.

All analyses were performed using R statistical language, Python and linux programming language on Terra (https://terra.bio/) and Verily workbench (https://workbench.verily.com/).

Our general workflow was illustrated in Figure 1.

## Results

### 1. Demographics

In the discovery and validation cohorts, the distributions of PD_*GBA1*_wildtype_ and PD_*GBA1*_ were similar between risk allele carriers and non-carrier groups across the lysosomal genetic variants ( *TMEM175* p.M393T, *TMEM175* p.Q65P, *SCARB2* rs6825004 and *CTSB* rs1293298) (Table S1-S2). In addition, the frequency of *GBA1* carriers was higher in the discovery cohort than in the validation cohort likely reflecting differences in cohort design, with *GBA1* carriers selectively included in the initial genotyping phase of the discovery cohort, while the validation cohort represents an unselected cohort.

In the discovery cohort, no significant differences in sex, disease onset, disease duration and age at genotyping were observed between risk and protective allele carriers for these five lysosomal genetic variants, after correction for multiple testing (*TMEM175*, *SCARB2* and *CTSB*) (Table 1 and Table S3).

**Table 1.**
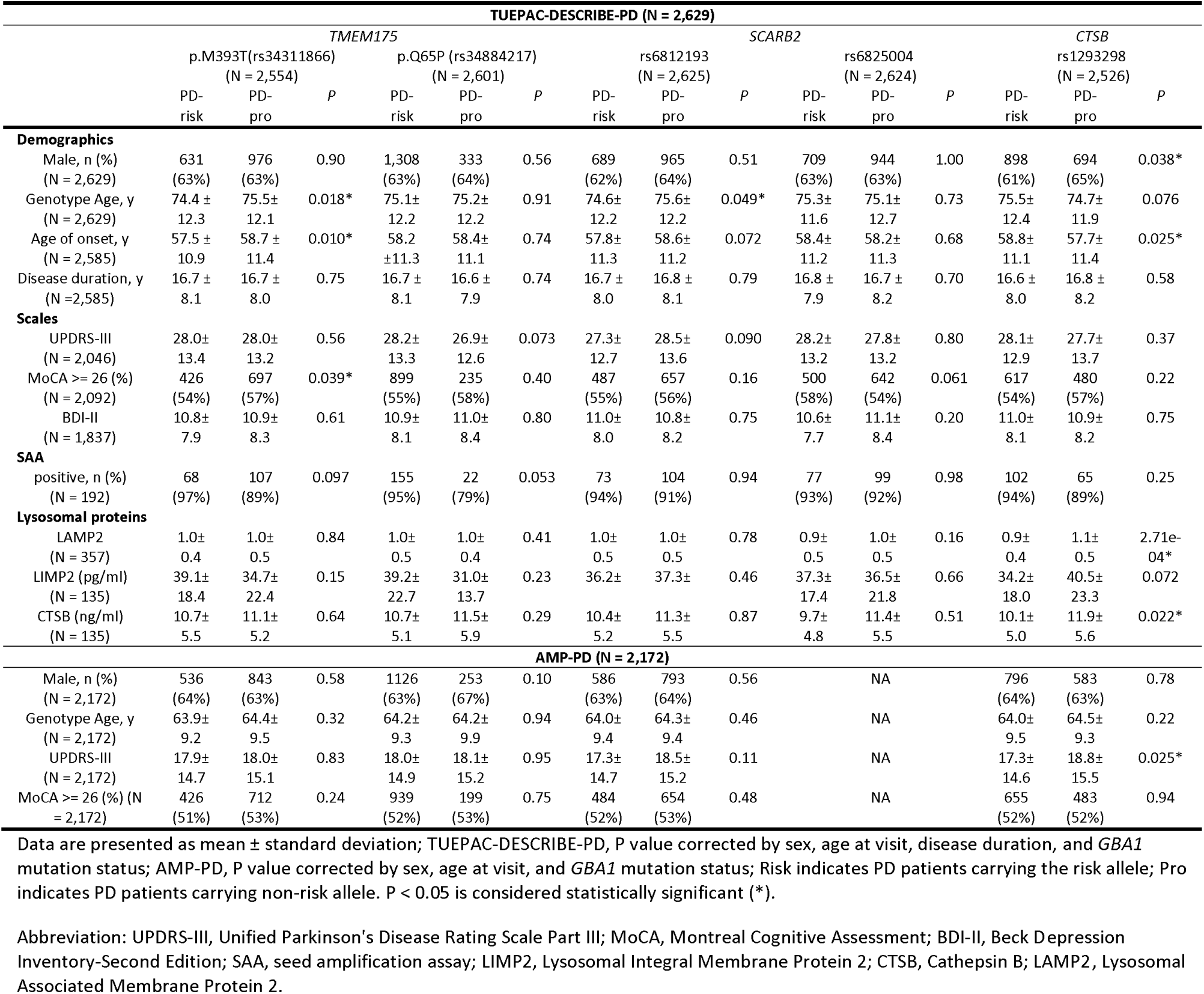
Comparison of demographics, clinical features, and lysosomal protein levels between risk allele carriers and protective allele carriers for genetic variants of three lysosome-related genes (*TMEM175*, *SCARB2*, and *CTSB*).

In the validation cohort, neither sex nor age for PD patients differed significantly between risk and protective allele carriers for all known genetic variants among lysosomal genes (*TMEM175*, *SCARB2*, and *CTSB*) (Table 1 and Table S3).

### 2. Clinical outcomes

In the discovery cohort, a slightly higher percentage of PD patients with MoCA >= 26 was observed among carriers of the *TMEM175* p.M393M protective allele compared with carriers of the *TMEM175* p.M393T risk allele (57% vs 54%, *P* = 0.039) in PD_all_ group (Table 1, Figure S1A). However, this did not remain significant after multiple-testing correction (FDR corrected *P* = 0.15) (Table S3). No significant differences were found between carriers of the p.M393T risk allele and the p.M393M protective allele in PD_*GBA1*_wildtype_ and PD_*GBA1*_ subgroups (Figure S1A, Figure S2A).

In addition, the difference between groups was not significant for UPDRS-III and BDI-II scores for all the examined genetic variants across lysosomal-related genes in the discovery cohort (*P* > 0.05) (Table 1 and Table S3).

In the validation cohort, no significant differences were observed in either the proportion of participants with MoCA >= 26 or continuous MoCA scores between carriers of the p.M393T risk allele and the p.M393M protective allele across PD_all_, PD_*GBA1*_wildtype_ and PD_*GBA1*_ groups (Figure S1B, Figure S2B). However, consistent with the result in the discovery cohort, the proportion of patients with MoCA >= 26 was also higher in carriers of the p.M393M protective allele than in carriers of the p.M393T risk allele in PD_all_ (53% vs 51%, *P* = 0.24, FDR corrected *P* = 0.94) (Table S3, Figure S1B).

Counterintuitively, carriers of *CTSB* risk allele had a slightly lower mean UPDRS-III score than carriers of *CTSB* protective allele (17.3 vs 18.8, *P* = 0.025) in the validation cohort (Table 1). This was not observed in the discovery cohort (28.1 vs 27.7, *P* = 0.37) (Table 1) and did not remain significant after multiple-testing correction (FDR corrected *P* = 0.10) in the validation cohort (Table S3).

### 3. a-Syn SAA

We assessed whether the proportion of PD patients with positive α-syn seeding activity differed between the carriers of risk and non-risk alleles across five genetic variants. In the discovery cohort, carriers of the protective *TMEM175* p.Q65P allele showed a less positive α-synuclein seeding activity than patients with the p.Q65Q risk allele (79% vs 95%, *P* = 0.053, FDR corrected *P* = 0.24) (Table 1 and Table S3). Additionally, no significant differences were observed for another variant *TMEM175* p.M393T (*P* = 0.33, FDR corrected *P* = 0.24) (Table 1 and Table S3). No data was available for the validation cohort. No significant differences were observed in the proportion of positive α-syn seeding activity across *SCARB2* and *CTSB* genotypes.

### 4. Lysosomal proteins

In the discovery cohort, CSF mean LAMP2 levels was significantly lower in carriers of *CTSB* risk allele compared with those carriers of *CTSB* protective allele (0.9 vs 1.1, *P* = 2.71e-04, FDR corrected *P* = 1.36e-03) in PD_all_ (Table 1 and Table S3). After stratification by *GBA1* subtypes, carriers of *CTSB* risk allele showed a significantly lower CSF mean LAMP2 levels compared with carriers of *CTSB* protective allele (0.94 vs 1.07, *P* = 9.01e-04, FDR corrected *P* = 4.51e-03) in PD_*GBA1*_wildtype_. However, in PD_*GBA1*_, LAMP2 was not significantly different between carriers of *CTSB* risk allele and *CTSB* protective allele (0.94 vs 1.15, *P* = 0.090, FDR corrected *P* = 0.23) (Figure 2A).

**Figure 2.**
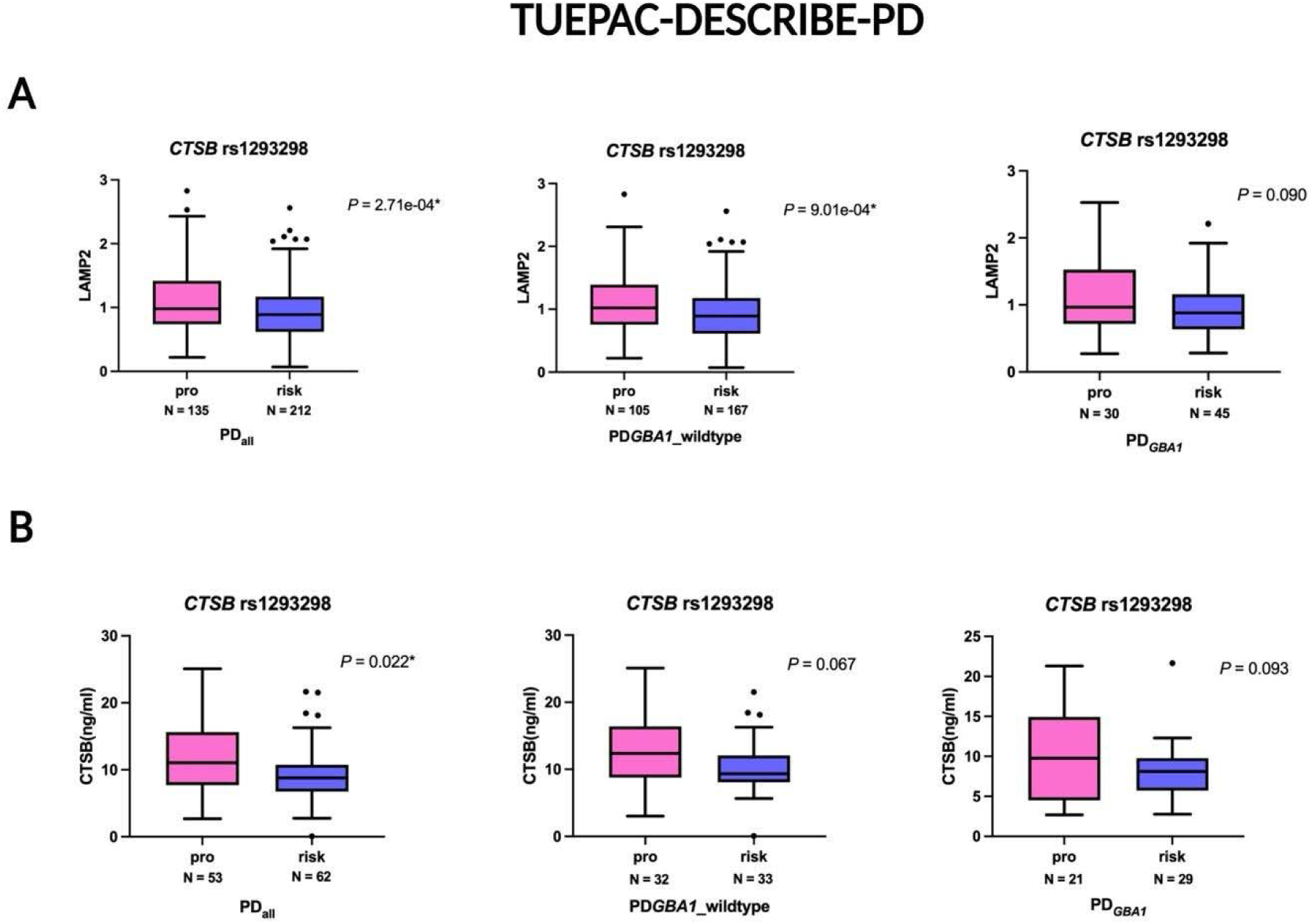
TUEPAC-DESCRIBE-PD. A) Lysosomal integral membrane protein 2 (LIMP2) and B) Cathepsin B(CTSB) levels, stratified by CTSB rs1293298 genotype (protective allele carriers [pro] vs. risk allele carriers [risk]) in PD_all_, PD_*GBA1*_wildtype_, and PD_*GBA1*_ subgroups [LAMP2(PD_all_:uncorrected P = 2.71e-04, FDR corrected P = 1.36e-03; PD_*GBA1*_wildtype_: uncorrected P = 9.01e-04, FDR corrected P = 4.51e-03), CTSB(PD_all_: uncorrected P = 0.022, FDR corrected P = 0.11; PD_*GBA1*_wildtype_: uncorrected P = 0.067, FDR corrected P = 0.34)].

In the discovery cohort, carriers of *CTSB* risk allele showed nominal significantly lower mean CSF CTSB compared with carriers of *CTSB* protective allele (10.1 vs 11.9 ng/ml, *P* = 0.022, FDR corrected *P* = 0.11) (Table 1 and Table S3). After stratification by *GBA1* subtypes, mean CSF levels of CTSB did not differ significantly between groups in PD_*GBA1*_wildtype_ (10.3 vs 12.5 ng/ml, *P* = 0.067, FDR corrected *P* = 0.34) (Figure 2B).

### 5. Sphingolipid profiles

In the discovery cohort, GCase activity in fresh blood leukocytes was nominal significantly lower in carriers of *SCARB2* rs6812193 risk allele compared to carriers of *SCARB2* rs6812193 protective allele (20.8% vs 23.7%, *P* = 0.033, FDR corrected *P* = 0.17) (Table 2 and Table S4). After stratification by *GBA1* subtypes, lysosomal GCase activity was significantly lower in carriers of *SCARB2* rs6812193 risk allele than carriers of *SCARB2* rs6812193 protective allele among PD_*GBA1*_wildtype_ patients (23.1% vs 29.0%, *P* = 5.31e-03, FDR corrected *P* = 0.027) in the discovery cohort (Figure 3A). No significant differences in GCase activity were observed between carriers of *SCARB2* rs6812193 risk allele and *SCARB2* rs6812193 protective allele within the PD_*GBA1*_ subgroup (19.9% vs 20.6%, *P* = 0.52, FDR corrected *P* = 0.85) (Figure 3A). In the discovery cohort, CSF levels of the GCase upstream substrates GlcCer C22:0, GlcCer C24:1, GlcCer C24:0 in CSF tended to be higher in carriers of the p.M393T risk allele compared with carriers of the p.M393M protective allele (GlcCer C22:0, 8.7 vs 7.4 nM, *P* = 0.25, FDR corrected *P* = 0.55; GlcCer C24:1, 1.8 vs 1.6 nM, *P* = 0.51, FDR corrected *P* = 0.64; GlcCer C24:0, 13.7 vs 12.0 nM, *P* = 0.36, FDR corrected *P* = 0.60). CSF level of GlcCer C24:0 was nominal significantly higher in carriers of *SCARB2* rs6825004 risk allele than carriers of *SCARB2* rs6825004 protective allele (14.0 vs 11.8, *P* = 0.034, FDR corrected *P* = 0.17). Carriers of the p.M393T risk allele showed nominal significantly lower CSF levels of the GCase downstream products Cer C16:0, Cer C18:0, Cer C20:0, Cer C22:0, and Cer C24:1 compared with carriers of the p.M393M protective allele in the discovery cohort (Cer C16:0, 1.8 vs 2.8 nM, *P* = 0.011, FDR corrected *P* = 0.055; Cer C18:0, 5.2 vs 6.5 nM, *P* = 0.034, FDR corrected *P* = 0.17; Cer C20:0, 0.6 vs 0.8 nM, *P* = 0.027, FDR corrected *P* = 0.14; Cer C22:0, 0.9 vs 1.3 nM, *P* = 0.019, FDR corrected *P* = 0.095; Cer C24:1, 2.9 vs 4.5 nM, *P* = 0.024, FDR corrected *P* = 0.12) (Table 2 and Table S4).

**Figure 3.**
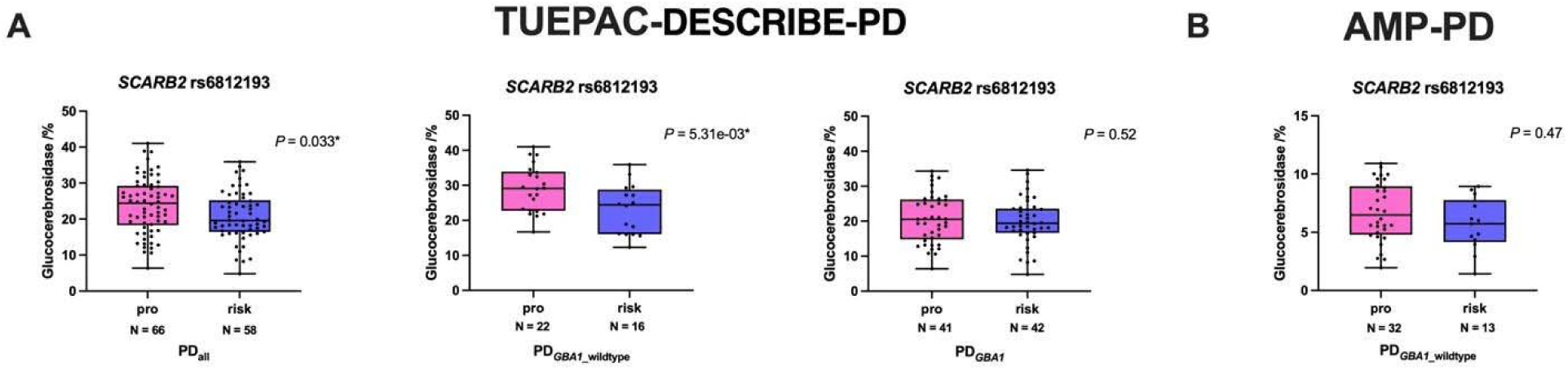
Associations between SCARB2 rs6812193 and Glucocerebrosidase (Gcase) in TUEPAC-DESCRIBE-PD and AMP-PD. A) TUEPAC-DESCRIBE-PD: Lysosomal GCase activity in protective(pro) and risk allele carriers of the SCARB2 rs6812193, stratified by GBA1: PD_*GBA1*_wildtype_, PD_*GBA1*_; Gcase (PD_all_:uncorrected P = 0.033, FDR corrected P = 0.17; PD_*GBA1*_wildtype_: uncorrected P = 5.31e-03, FDR corrected P = 0.027). B) AMP-PD: Lysosomal GCase activity in protective(pro) and risk allele carriers of the SCARB2 rs6812193 among GBA1 wildtype individuals. Data for GBA1 carriers in AMP-PD were not shown due to the limited number of cases (three patients with PD_*GBA1*_risk_ and one patient with PD_*GBA1*_mild_). Gcase (PD_*GBA1*_wildtype_: uncorrected P = 0.27, FDR corrected P = 0.36).

**Table 2.**
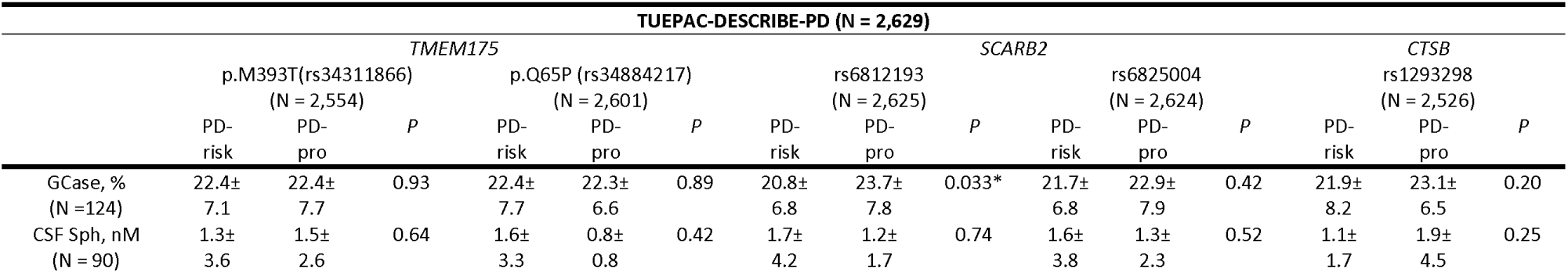
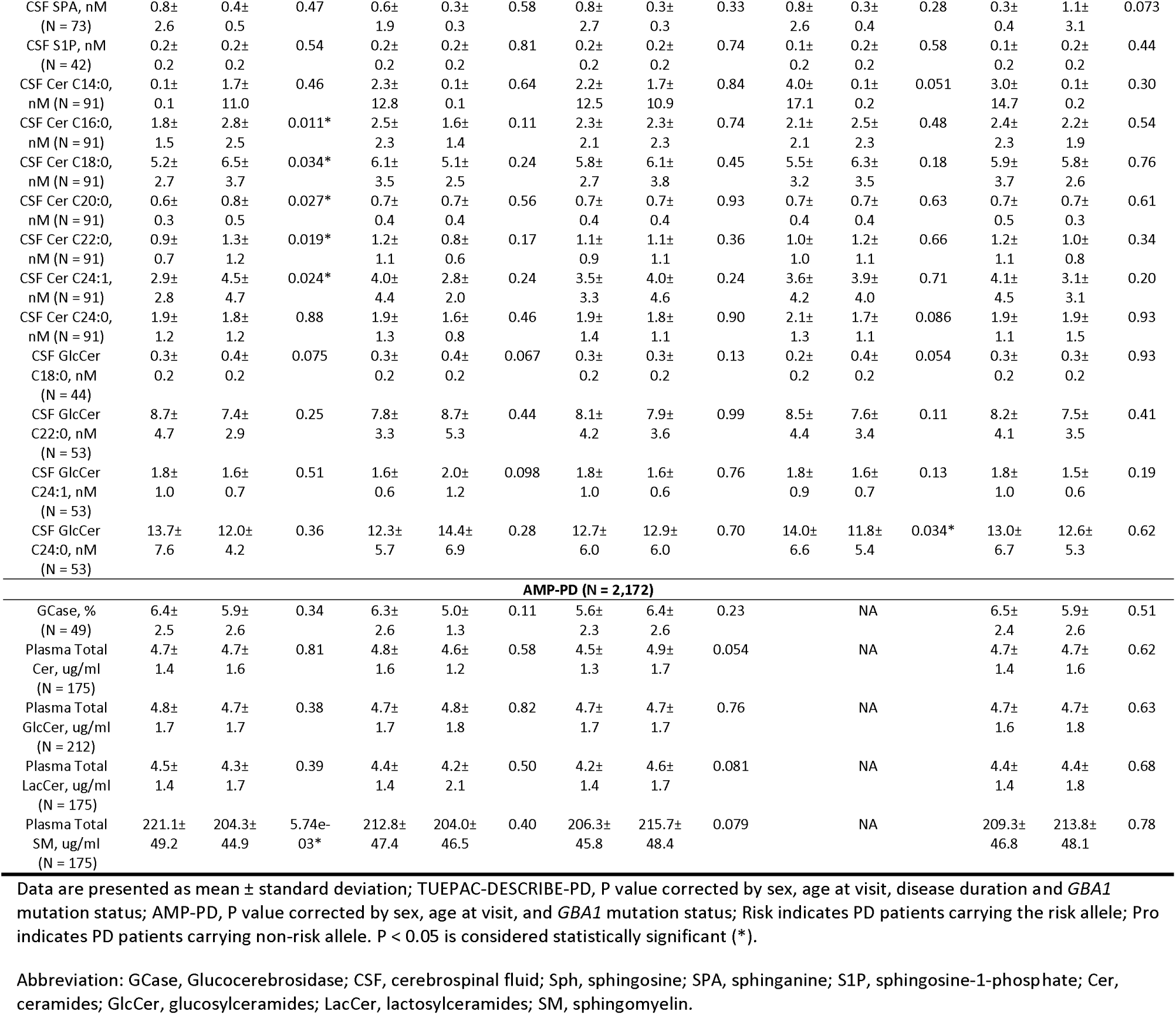
CSF and plasma levels of sphingolipids profiles in TUEPAC-DESCRIBE-PD and AMP-PD cross-sectionally.

In the validation cohort, carriers of *SCARB2* rs6812193 risk allele also showed a trend toward reduced CSF GCase activity compared to carriers of *SCARB2* rs6812193 protective allele; however, this difference did not reach statistical significance (5.6% vs 6.4%, *P* = 0.47, FDR corrected *P* = 0.56) (Table 2 and Table S4). In PD_*GBA1*_wildtype_, carriers of *SCARB2* rs6812193 risk allele also showed lower GCase activity compared with carriers of *SCARB2* rs6812193 protective allele in the validation cohort, although this difference did not reach statistical significance (5.6% vs. 6.7%, *P* = 0.27, FDR corrected *P* = 0.36) (Figure 4B). Analyses in PD_*GBA1*_ subgroups could not be performed due to the limited number of cases. In addition, SM, which is hydrolyzed by sphingomyelinase to generate ceramide, was significantly higher in carriers of the p.M393T risk allele than carriers of the p.M393M protective allele (221.1 vs 204.3, *P* = 5.74e-03, FDR corrected *P* = 0.023) (Table 2 and Table S4).

### 6. Associations of UPDRS-III, MoCA, BDI-I I, lysosomal proteins with Sphingolipid profiles

In the discovery cohort, there were no significant associations between clinical outcomes (UPDRS-III, MoCA and BDI-II) and sphingolipid profiles (Table S5); higher LAMP2 levels were significantly associated with lower UPDRS-III score (rho = −0.14, FDR corrected *P* = 0.034), lower ceramides levels (Cer C22:0: rho = −0.29, FDR corrected *P* = 0.044; Cer C24:1: rho = −0.39, FDR corrected *P* = 2.85e-03; Cer C24:0: rho = - 0.43, FDR corrected *P* = 5.87e-04) and lower glycosylceramides levels (GlcCer C22:0: rho = −0.52, FDR corrected *P* = 1.25e-03; GlcCer C24:1: rho = −0.52, FDR corrected *P* = 1.52e-03; GlcCer C24:0: rho = −0.44, FDR corrected *P* =0.010) (Table S6).

## Discussion

In PD patients, our data revealed associations between comprehensive dimensions of biomarker profiles, clinical features and genetic variants across three lysosomal-related genes (*TMEM175*, *SCARB2*, and *CTSB*). Specifically, *TMEM175* p.M393T risk variant was nominally associated with worse cognitive performance in the discovery cohort. *TMEM175* p.Q65P risk allele carriers were shown to have a higher percentage of positive α-syn seeding activity compared to non-carriers, although this difference did not reach statistical significance. Moreover, *SCARB2* rs6812193 risk allele was associated with reduced GCase activity. Additionally, *CTSB* rs1293298 risk variant are linked to decreased CSF levels of lysosomal proteins (CTSB, and LAMP2). Finally, nominal differences in sphingolipid profiles were observed in PD patients carrying *TMEM175* p.M393T variants.

Consistent with previous large-scale studies^9, 12^, p.M393T risk allele in *TMEM175* was nominally associated with an earlier age at onset in 2,629 PD patients of the discovery cohort. By contrast, the protective allele (C) of *CTSB* rs1293298 was unexpectedly associated with an earlier age at onset in the discovery cohort. This nominal finding contrasts with the age-of-onset GWAS by Blauwendraat et al.^9^, which reported that the C allele is associated with a modestly later onset (β = +0.218 years per allele). Such apparent discrepancies may reflect limited statistical power in a regional cohort, differences in allele frequency, environmental and recruitment-related factors.

We did not observe an association between the *TMEM175* p.M393T variant and motor severity in either the discovery cohort or the validation cohort (N > 2,000), in contrast to a previous smaller study^8^. This discrepancy may reflect limited statistical power in that cohort of 336 patients, as well as potential regional differences. Regarding *CTSB* rs1293298, no difference in UPDRS-III scores was observed in the discovery cohort, whereas in the validation cohort risk allele carriers showed slightly lower scores, an unexpected result given the known risk association of this locus. In the validation cohort, patient had lower UPDRS-III scores compared to the discovery cohort, which might reflect a different recruitment strategy. The validation cohort included a subset of PD patients recruited at early stage, whereas the discovery cohort consisted of patients followed annually in outpatient clinics. We therefore hypothesized that the genetic effects are more pronounced in early stages of disease. Together, these factors may obscure or even yield counterintuitive genotype–phenotype associations.

Our results were consistent with a previous report^8^, which showed that PD patients carrying the *TMEM175* p.M393T risk variant exhibited worse cognitive performance in the discovery cohort when *GBA1* status was included as a covariate, given the established impact of *GBA1* on cognition in PD^40–43^. However, cognition in PD was also affected by covariates such as age at visit, disease duration, and sex. These factors may confound genotype–phenotype associations. Importantly, this association did not survive correction for multiple testing and should therefore be interpreted with caution. To further minimize *GBA1* effects, we performed a sensitivity analysis excluding *GBA1* carriers; the direction of effect was similar, with p.M393T carriers showing a higher prevalence of cognitive impairment. These findings suggested a potential role of p.M393T as a genetic contributor to cognitive impairment in PD, although this remained exploratory and required confirmation in independent cohorts. Interestingly, the channel encoded by *TMEM175* is thought to be druggable, and currently ongoing work is exploring pharmacological strategies to modulate its activity^44–46^. Future clinical trials evaluating ion channel activators in PD patients may consider incorporating cognitive measures such as the MoCA score as exploratory markers to monitor treatment efficacy, offering a promising avenue for therapeutic development.

In the discovery cohort, PD patients carrying the *TMEM175* p.Q65P non-risk allele showed a less α-syn seeding activity than risk allele carriers. However, this difference did not reach statistical significance, likely due to the limited number of PD patients with negative seeding. Larger powered studies are needed to validate this association. If confirmed, future clinical trials targeting α-syn pathology could incorporate genetic stratification (e.g., p.Q65P) to enhance patient selection and facilitate monitoring of treatment response.

In line with a previous study^47^, which reported no effect of the two known *SCARB2* variants on LIMP2 expression in controls, our findings extended this observation to PD patients, suggesting that risk variants in *SCARB2* were unlikely to contribute to PD pathogenesis through the modulation of LIMP2 expression. In contrast to *SCARB2*, we observed nominally decreased CTSB expression in carriers of *CTSB* rs1293298 risk allele, which may indicate impaired lysosomal proteolytic activity. This was consistent with previous studies linking the risk variant to lysosomal dysfunction^20^. Given that CTSB is involved in the lysosomal degradation of α-syn^21^, reduced CTSB expression could potentially impair α-syn clearance. On the other hand, we excluded *GBA1* carriers to avoid potential bias due to *GBA1*-related lysosomal effects. The difference in CTSB protein levels between genotype groups was no longer significant. This may suggest that the loss of significance for CTSB protein levels could be due to limited statistical power after stratification, given the smaller sample size in the non-*GBA1* subgroup.

In addition, we identified an association between *CTSB* rs1293298 and reduced LAMP2 protein expression in the discovery cohort. Although a direct link between *CTSB* and LAMP2 was not previously reported in PD patients, inhibition of CTSB has been shown to promote lysosomal dysfunction and increase LAMP2 immunofluorescence in cell models^20^. In the current study, PD patients carrying the *CTSB* rs1293298 risk allele showed decreased CSF LAMP2 expression compared with non-carriers. This observation may be biologically relevant, as LAMP2 compromises a major protein component of lysosomal membranes and plays critical roles in both autophagy and chaperone-mediated autophagy (CMA)^48^. Reduced LAMP2 levels have also been reported in CSF^31^, brain tissue^49^ and peripheral leukocytes^50^ of patients with α-synucleinopathies, including PD and Lewy body dementia. Taken together, our results supported the notion that *CTSB* risk variants impaired hydrolase activity, further exacerbate α-syn aggregation and lysosomal dysfunction, with abnormal LAMP2 expression as a downstream consequence. These findings warrant replication in independent cohorts and further experimental studies to clarify the mechanistic relationship between *CTSB* variant and LAMP2 regulation in PD.

Interestingly, we also observed significant correlations between LAMP2 protein levels and sphingolipid profiles in PD. Reduced LAMP2 may contribute to impaired autophagy in PD, in turn, to disturbances in lipid metabolism^51^. Conversely, sphingolipid has been reported to regulate LAMP2^52^. Together, these findings further highlight the important interactions between sphingolipid and LAMP2 mediated autophagy in PD.

*TMEM175* p.M393T has previously been reported to be associated with reduced GCase activity^11^, but we did not replicate this finding, as no difference in GCase activity was observed in either of the cohorts analyzed. In contrast, we found significantly reduced GCase activity among *SCARB2* rs6812193 risk allele carriers compared with protective carriers in the discovery cohort, with a similar but non-significant trend observed in the validation cohort after excluding *GBA1* carriers. The use of distinct biomaterials and analytical methods to measure GCase activity may explain the inconsistent results observed across cohorts. Additionally, these findings differ from a previous study^53^, which may reflect methodological differences, regional cohort bias or limited statistical power, as the GCase dataset in our cohorts was relatively smaller and may still be underpowered. From a biological perspective, *SCARB2* encodes the lysosomal integral membrane protein (LIMP2), which functions as the receptor responsible for transporting GCase to lysosomes. Thus, impairment of LIMP2 function could directly influence GCase activity and contribute to PD pathogenesis.

Comprehensive sphingolipid profiling in the discovery cohort and the validation cohorts revealed distinct alterations across different steps of the lysosomal pathway. (1) No significant differences in CSF levels of GCase upstream substrates (glucosylceramides) were observed for *TMEM175* p.M393T. (2) CSF levels of the GCase downstream product ceramides were nominally reduced in PD patients with *TMEM175* p.M393T risk variant compared with non-carriers; (3) Plasma levels of the by-product sphingomyelin were higher in PD patients with *TMEM175* p.M393T risk allele compared with non-carriers. Together, these findings implicate p.M393T risk variant in abnormal regulation of sphingolipid metabolism in biofluids.

Mechanistically, GCase hydrolyzes glucosylceramides and glucosylsphingosines to generate ceramides, sphingosine, and sphingosine-1-phosphate^1^. Therefore, GCase deficiency leads to accumulation of upstream substrates (glucosylceramides), while ceramide production is diminished. In addition, *SMPD1* encodes the lysosomal enzyme acid sphingomyelinase (ASMase), and genetic variants in *SMPD1* has been associated with increased risk of PD^54–56^. ASMase hydrolyzes sphingomyelin, a major membrane lipid, to generate ceramide and thereby contributes to the ceramide pool. Thus, *SMPD1*/ASMase may represent another lysosomal pathway through which altered sphingolipid metabolism contributes to PD pathogenesis. The *TMEM175* p.M393T variant may also influence enzymes involved in other lysosomal pathways, ultimately resulting in accumulation of sphingomyelin and reduced ceramide production beyond the GCase–glucosylceramide–ceramide axis. Overall, our results suggested that *TMEM175* risk variant contribute to impaired lysosomal enzyme activity, upstream substrate accumulation, and reduced downstream ceramide production, leading to broader defects in lipid metabolism and lysosomal function in PD.

Importantly, the biochemical assessments of GCase activity and sphingolipid species highlight how genetic variants in three lysosomal-related genes studied manifest at a functional level in patient-derived biofluids. This provides insights that are often missing when translating findings from cell or animal models into patient cohorts. Future work should determine whether these biochemical measures could serve as read-outs for target engagement in upcoming clinical trials.

A strength of the present study is that we validated part of the findings from the TUEPAC-DESCRIBE-PD cohort in the AMP-PD cohort. The relatively large sample size and combined genotyping of lysosomal genetic variants together with *GBA1* screening enabled gene-specific analyses. Furthermore, this is the first study to integrate these lysosomal genetic variants, which seem to closely interact as previously highlighted in GWAS, with a broad range of biomarkers, including clinical outcomes, α-syn seeding activity, lysosomal protein expression, and sphingolipid profiles.

There are several limitations. Accurate detection of *GBA1* variants from short-read WGS remains challenging because of the high sequence homology between *GBA1* and its adjacent pseudogene, *GBAP1*. Therefore, some complex, recombinant, or rare variants may not have been reliably detected. Moreover, age at onset was unavailable in the validation dataset, precluding calculation of disease duration; as a result, AMP-PD analyses were not adjusted for disease duration, potentially influencing the observed associations. These limitations should be considered when interpreting the findings. Finally, as this is an exploratory study, not all measures could be replicated in AMP-PD, and some associations were nominal and did not survive after multiple testing correction.

In conclusion, we identified associations between lysosomal genetic variants and clinical or fluid biomarker profiles in PD, including *TMEM175* p.M393T with cognition and sphingolipid profiles, *CTSB* rs1293298 with lysosomal protein levels, and *SCARB2* rs6812193 with GCase activity. Although the observed effects were modest and some associations did not survive correction for multiple testing, these findings suggested that lysosomal genetic variants may contribute to biological heterogeneity in PD. They may therefore provide useful exploratory reference data for future clinical trials focused on patient stratification, biomarker selection, and lysosomal pathway-targeted therapies.

## Data and Code Availability Statement

Data used in the preparation of this article were obtained from the Global Parkinson’s Genetics Program (GP2; https://gp2.org) and the Accelerating Medicine Partnership® (AMP®) Parkinson’s Disease (AMP PD) Knowledge Platform(https://amp-pd.org). Specifically, we used Tier 2 data from GP2 release 7 (https://zenodo.org/records/10962119) and AMP-PD release 4 (https://www.amp-pd.org/news/gp2-release-notes-november-2023). GP2 data are available on AMP PD (https://amp-pd.org).

All code generated for this article, and the identifiers for all software programs and packages used, are available on GitHub (https://github.com/wenhuasun/TMEM175_Biomarker/tree/main).

## Acknowledgements

This project was supported by the Global Parkinson’s Genetics Program (GP2; https://gp2.org). GP2 is funded by the Aligning Science Across Parkinson’s (ASAP) initiative and implemented by The Michael J. Fox Foundation for Parkinson’s Research (MJFF). For the purpose of Open Access, the author has applied a CC BY public copyright licence to any Author Accepted Manuscript version arising from this submission.

Data used in the preparation of this article were obtained from the Accelerating Medicine Partnership® (AMP®) Parkinson’s Disease (AMP PD) Knowledge Platform. For up-to-date information on the study, visit https://www.amp-pd.org.

The AMP® PD program is a public-private partnership managed by the Foundation for the National Institutes of Health and funded by the National Institute of Neurological Disorders and Stroke (NINDS) in partnership with the Aligning Science Across Parkinson’s (ASAP) initiative; Celgene Corporation, a subsidiary of Bristol-Myers Squibb Company; GlaxoSmithKline plc (GSK); The Michael J. Fox Foundation for Parkinson’s Research; AbbVie Inc.; Pfizer Inc.; Sanofi US Services Inc.; and Verily Life Sciences.

ACCELERATING MEDICINES PARTNERSHIP and AMP are registered service marks of the U.S. Department of Health and Human Services.

This work was conducted as part of a project supported by the Michael J. Fox Foundation for Parkinson’s Research (MJFF).

W.S. acknowledges the support of the China Scholarship Council program (Project ID: 202207040033).

H.Z. is a Wallenberg Scholar and a Distinguished Professor at the Swedish Research Council supported by grants from the Swedish Research Council (#2023-00356, #2022-01018 and #2019-02397), the European Union’s Horizon Europe research and innovation programme under grant agreement No 101053962, Swedish State Support for Clinical Research (#ALFGBG-71320)

## Contributions

(1) Research project: A. Conception, B. Organization, C. Execution; (2) Statistical analysis: A. Design, B. Execution, C. Review and critique; (3) Manuscript preparation: A. Writing of the first draft, B. Review and critique.

W.S.: 1A, 1B, 1C, 2A, 2B, 2C, 3A, 3B

I.W.: 1C, 3B

B.R.: 1C, 3B

R.K.: 1C, 3B

M.M.: 1C, 3B

H.Z.: 1C, 3B

S.L.: 1A, 1B, 1C, 3B

A.K.H.: 1C, 3B

C.S.: 1A, 1B, 1C, 2C, 3B

P.P.: 1C, 3B

G.C.P.: 3B

A.S.: 3B

U.W.: 3B

C.v.R.: 3B

F.M.: 3B

B.F.: 3B

B.M.: 3B

I.Z.: 3B

E.D.: 3B

P.L.: 3B

A.W.: 3B

J.L.: 3B

W.H.: 3B

M.L.: 3B

Z.G.O.: 1A,3B

K.B.: 1A, 3B

T.G.: 1A, 1B, 1C, 3B

