## Supplementary figures and tables for "Stratified cohorts for biomarker assessment and trial readiness: *TMEM175*, *SCARB2* and *CTSB* in Parkinson’s disease"

Table S1. Distribution of *GBA1* subtypes across genetic variants of lysosomal genes (*TMEM175*, *SCARB2*, and *CTSB*) in TUEPAC-DESCRIBE-PD.

| Genotypes | | PD*_GBA1_*__wildtype_ | PD*_GBA1_* | Total | P value | FDR corrected *P* value |
| --- | --- | --- | --- | --- | --- | --- |
| *TMEM175* p.M393T  (risk allele freq(C)=0.22) | risk (CC/CT) | 826 (83%) | 166 (17%) | 992 | 0.51 | 0.58 |
|  | pro (TT) | 1,278(84%) | 239(16%) | 1,517 |  |  |
| *TMEM175* p.Q65P  (risk allele freq(A)=0.90) | risk (AA) | 1,707(83%) | 343(17%) | 2,050 | 0.074 | 0.19 |
|  | pro (CC/CA) | 437(87%) | 68(13%) | 505 |  |  |
| *SCARB2* rs6812193  (Risk allele freq(C)=0.65) | risk (CC) | 895(82%) | 194(18%) | 1,089 | 0.029* | 0.15 |
|  | pro (CT/TT ) | 1,272(85%) | 218(15%) | 1,490 |  |  |
| *SCARB2* rs6825004  (Risk allele freq(C)=0.65) | risk (CC) | 941(85%) | 169(15%) | 1,110 | 0.39 | 0.58 |
|  | pro (CG/GG) | 1,226 (84%) | 242 (16%) | 1,468 |  |  |
| *CTSB* rs1293298  (Risk allele freq(A)=0.76) | risk (AA) | 1,206 (84%) | 223(16%) | 1,436 | 0.58 | 0.58 |
|  | pro (CA/CC) | 887(85%) | 159(15%) | 1,046 |  |  |

P value represents the chi-square statistic to assess the association between any *GBA1* mutation status (wildtype vs. *GBA1* carriers) and the genetic variants. P values were corrected for multiple testing across five genetic variants using the Benjamini–Hochberg method to control the false discovery rate.

| Genotypes | | PD*_GBA1_*__wildtype_ | PD*_GBA1_* | Total | P value | FDR corrected *P* value |
| --- | --- | --- | --- | --- | --- | --- |
| *TMEM175* p.M393T  (Risk allele freq(C)=0.22) | risk (CC/CT) | 764(92%) | 70(8%) | 834 | 0.87 | 0.87 |
|  | pro (TT) | 1,223 (91%) | 115(9%) | 1338 |  |  |
| *TMEM175* p.Q65P  (risk allele freq(A)=0.91) | risk (AA) | 1,640(91%) | 156(9%) | 1,796 | 0.54 | 0.72 |
|  | pro (CC/CA) | 347(92%) | 29(8%) | 376 |  |  |
| *SCARB2* rs6812193  (Risk allele freq(C)=0.65) | risk (CC) | 864(93%) | 70(7%) | 934 | 0.14 | 0.56 |
|  | pro (CT/TT) | 1,123(91%) | 115(9%) | 1,238 |  |  |
| *CTSB* rs1293298  (Risk allele freq(A)=0.76) | risk (AA) | 1,136(91%) | 112(9%) | 1,248 | 0.38 | 0.72 |
|  | pro (CA/CC) | 851 (92%) | 73(8%) | 924 |  |  |

Table S2. Distribution of *GBA1* subtypes across genetic variants of lysosomal genes (*TMEM175*, *SCARB2*, and *CTSB*) in AMP-PD.

P value represents the chi-square statistic to assess the association between any *GBA1* mutation status (wildtype vs. *GBA1* carriers) and the genetic variants. P values were corrected for multiple testing across four genetic variants using the Benjamini–Hochberg method to control the false discovery rate.

Table S3. Comparison of demographics, clinical features, and lysosomal protein levels between risk allele carriers and protective allele carriers for genetic variants of three lysosome-related genes (*TMEM175*, *SCARB2*, and *CTSB*), after correction for multiple testing.

| **TUEPAC-DESCRIBE-PD (N = 2,629)** | | | | | | | | | | | | | | | |
| --- | --- | --- | --- | --- | --- | --- | --- | --- | --- | --- | --- | --- | --- | --- | --- |
|  | *TMEM175* | | | | | | *SCARB2* | | | | | | *CTSB* | | |
|  | p.M393T(rs34311866)  (N = 2,554) | | | p.Q65P (rs34884217)  (N = 2,601) | | | rs6812193  (N = 2,625) | | | rs6825004  (N = 2,624) | | | rs1293298   (N = 2,526) | | |
|  | PD-risk | PD-pro | FDR corrected *P* | PD-risk | PD-pro | FDR corrected *P* | PD-risk | PD-pro | FDR corrected *P* | PD-risk | PD-pro | FDR corrected *P* | PD-risk | PD-pro | FDR corrected *P* |
| **Demographics** | | | | | | | | | | | | | | | |
| Male, n (%)  (N = 2,629) | 631  (63%) | 976  (63%) | 1.00 | 1,308  (63%) | 333  (64%) | 0.93 | 689  (62%) | 965  (64%) | 0.93 | 709  (63%) | 944  (63%) | 1.00 | 898  (61%) | 694  (65%) | 0.19 |
| Genotype Age, y  (N = 2,629) | 74.4 ±  12.3 | 75.5±  12.1 | 0.090 | 75.1±  12.2 | 75.2±  12.2 | 0.91 | 74.6±  12.2 | 75.6±  12.2 | 0.12 | 75.3±  11.6 | 75.1±  12.7 | 0.91 | 75.5±  12.4 | 74.7±  11.9 | 0.13 |
| Age of onset, y  (N = 2,585) | 57.5 ±  10.9 | 58.7 ±  11.4 | 0.05 | 58.2  ±11.3 | 58.4±  11.1 | 0.74 | 57.8±  11.3 | 58.6±  11.2 | 0.12 | 58.4±  11.2 | 58.2±  11.3 | 0.74 | 58.8±  11.1 | 57.7±  11.4 | 0.063 |
| Disease duration, y (N =2,585) | 16.7 ±  8.1 | 16.7 ±  8.0 | 0.79 | 16.7 ±  8.1 | 16.6 ±  7.9 | 0.79 | 16.7 ±  8.0 | 16.8 ±  8.1 | 0.79 | 16.8 ±  7.9 | 16.7 ±  8.2 | 0.79 | 16.6 ±  8.0 | 16.8 ±  8.2 | 0.79 |
| **Scales** | | | | | | | | | | | | | | | |
| UPDRS-III  (N = 2,046) | 28.0±  13.4 | 28.0±  13.2 | 0.70 | 28.2±  13.3 | 26.9±  12.6 | 0.23 | 27.3±  12.7 | 28.5±  13.6 | 0.23 | 28.2±  13.2 | 27.8±  13.2 | 0.80 | 28.1±  12.9 | 27.7±  13.7 | 0.62 |
| MoCA >= 26 (%)  (N = 2,092) | 426  (54%) | 697  (57%) | 0.15 | 899  (55%) | 235  (58%) | 0.40 | 487  (55%) | 657  (56%) | 0.27 | 500  (58%) | 642  (54%) | 0.15 | 617  (54%) | 480  (57%) | 0.28 |
| BDI-II  (N = 1,837) | 10.8±  7.9 | 10.9±  8.3 | 0.80 | 10.9±  8.1 | 11.0±  8.4 | 0.80 | 11.0±  8.0 | 10.8±  8.2 | 0.80 | 10.6±  7.7 | 11.1±  8.4 | 0.80 | 11.0±  8.1 | 10.9±  8.2 | 0.80 |
| **SAA** | | | | | | | | | | | | | | | |
| positive, n (%)  (N = 192) | 68  (97%) | 107  (89%) | 0.24 | 155  (95%) | 22  (79%) | 0.24 | 73  (94%) | 104  (91%) | 0.98 | 77  (93%) | 99  (92%) | 0.98 | 102  (94%) | 65  (89%) | 0.42 |
| **Lysosomal proteins** | | | | | | | | | | | | | | | |
| LAMP2  (N = 357) | 1.0±  0.4 | 1.0±  0.5 | 0.84 | 1.0±  0.5 | 1.0±  0.4 | 0.68 | 1.0±  0.5 | 1.0±  0.5 | 0.84 | 0.9±  0.5 | 1.0±  0.5 | 0.40 | 0.9±  0.4 | 1.1±  0.5 | 1.36e-03* |
| LIMP2 (pg/ml)  (N = 135) | 39.1±  18.4 | 34.7±  22.4 | 0.38 | 39.2±  22.7 | 31.0±  13.7 | 0.38 | 36.2± | 37.3± | 0.58 | 37.3±  17.4 | 36.5±  21.8 | 0.66 | 34.2±  18.0 | 40.5±  23.3 | 0.36 |
| CTSB (ng/ml)  (N = 135) | 10.7±  5.5 | 11.1±  5.2 | 0.80 | 10.7±  5.1 | 11.5±  5.9 | 0.73 | 10.4±  5.2 | 11.3±  5.5 | 0.87 | 9.7±  4.8 | 11.4±  5.5 | 0.80 | 10.1±  5.0 | 11.9±  5.6 | 0.11 |
| **AMP-PD (N = 2,172)** | | | | | | | | | | | | | | | |
| Male, n (%)  (N = 2,172) | 536  (64%) | 843  (63%) | 0.77 | 1126  (63%) | 253  (67%) | 0.40 | 586  (63%) | 793  (64%) | 0.77 | NA | | | 796  (64%) | 583  (63%) | 0.78 |
| Genotype Age, y  (N = 2,172) | 63.9±  9.2 | 64.4±  9.5 | 0.61 | 64.2±  9.3 | 64.2±  9.9 | 0.94 | 64.0±  9.4 | 64.3±  9.4 | 0.61 | NA | | | 64.0±  9.5 | 64.5±  9.3 | 0.61 |
| UPDRS-III  (N = 2,172) | 17.9±  14.7 | 18.0±  15.1 | 0.95 | 18.0±  14.9 | 18.1±  15.2 | 0.95 | 17.3±  14.7 | 18.5±  15.2 | 0.22 | NA | | | 17.3±  14.6 | 18.8±  15.5 | 0.10 |
| MoCA >= 26 (%)  (N = 2,172) | 426  (51%) | 712  (53%) | 0.94 | 939  (52%) | 199  (53%) | 0.94 | 484  (52%) | 654  (53%) | 0.94 | NA | | | 655  (52%) | 483  (52%) | 0.94 |

Data are presented as mean ± standard deviation; TUEPAC-DESCRIBE-PD, P value corrected by sex, age at visit, disease duration, and *GBA1* mutation status; AMP-PD, P value corrected by sex, age at visit, and *GBA1* mutation status; Risk indicates PD patients carrying the risk allele; Pro indicates PD patients carrying non-risk allele. P < 0.05 is considered statistically significant (*). P values were corrected for multiple testing across five genetic variants using the Benjamini–Hochberg method to control the false discovery rate.

Abbreviation: UPDRS-III, Unified Parkinson's Disease Rating Scale Part III; MoCA, Montreal Cognitive Assessment; BDI-II, Beck Depression Inventory-Second Edition; SAA, seed amplification assay; LIMP2, Lysosomal Integral Membrane Protein 2; CTSB, Cathepsin B; LAMP2, Lysosomal Associated Membrane Protein 2.

Table S4. CSF and plasma levels of sphingolipids profiles in TUEPAC-DESCRIBE-PD and AMP-PD cross-sectionally, after correction for multiple testing.

| **TUEPAC-DESCRIBE-PD (N = 2,629)** | | | | | | | | | | | | | | | | |
| --- | --- | --- | --- | --- | --- | --- | --- | --- | --- | --- | --- | --- | --- | --- | --- | --- |
|  | *TMEM175* | | | | | | *SCARB2* | | | | | | | *CTSB* | | |
|  | p.M393T(rs34311866)  (N = 2,554) | | | p.Q65P (rs34884217)  (N = 2,601) | | | rs6812193  (N = 2,625) | | | rs6825004  (N = 2,624) | | | | rs1293298  (N = 2,526) | | |
|  | PD-risk | PD-pro | FDR corrected *P* | PD-risk | PD-pro | FDR corrected  *P* | PD-risk | PD-pro | FDR corrected *P* | | PD-risk | PD-pro | FDR corrected *P* | PD-risk | PD-pro | FDR corrected *P* |
| GCase, %  (N =124) | 22.4±  7.1 | 22.4±  7.7 | 0.93 | 22.4±  7.7 | 22.3±  6.6 | 0.93 | 20.8±  6.8 | 23.7±  7.8 | 0.17 | | 21.7±  6.8 | 22.9±  7.9 | 0.70 | 21.9±  8.2 | 23.1±  6.5 | 0.50 |
| CSF Sph, nM  (N = 90) | 1.3±  3.6 | 1.5±  2.6 | 0.74 | 1.6±  3.3 | 0.8±  0.8 | 0.74 | 1.7±  4.2 | 1.2±  1.7 | 0.74 | | 1.6±  3.8 | 1.3±  2.3 | 0.74 | 1.1±  1.7 | 1.9±  4.5 | 0.74 |
| CSF SPA, nM  (N = 73) | 0.8±  2.6 | 0.4±  0.5 | 0.58 | 0.6±  1.9 | 0.3±  0.3 | 0.58 | 0.8±  2.7 | 0.3±  0.3 | 0.55 | | 0.8±  2.6 | 0.3±  0.4 | 0.55 | 0.3±  0.4 | 1.1±  3.1 | 0.37 |
| CSF S1P, nM  (N = 42) | 0.2±  0.2 | 0.2±  0.2 | 0.81 | 0.2±  0.2 | 0.2±  0.2 | 0.81 | 0.2±  0.2 | 0.2±  0.2 | 0.81 | | 0.1±  0.2 | 0.2±  0.2 | 0.81 | 0.1±  0.2 | 0.2±  0.2 | 0.81 |
| CSF Cer C14:0, nM  (N = 91) | 0.1±  0.1 | 1.7±  11.0 | 0.77 | 2.3±  12.8 | 0.1±  0.1 | 0.80 | 2.2±  12.5 | 1.7±  10.9 | 0.84 | | 4.0±  17.1 | 0.1±  0.2 | 0.26 | 3.0±  14.7 | 0.1±  0.2 | 0.75 |
| CSF Cer C16:0, nM  (N = 91) | 1.8±  1.5 | 2.8±  2.5 | 0.055 | 2.5±  2.3 | 1.6±  1.4 | 0.28 | 2.3±  2.1 | 2.3±  2.3 | 0.74 | | 2.1±  2.1 | 2.5±  2.3 | 0.68 | 2.4±  2.3 | 2.2±  1.9 | 0.68 |
| CSF Cer C18:0, nM  (N = 91) | 5.2±  2.7 | 6.5±  3.7 | 0.17 | 6.1±  3.5 | 5.1±  2.5 | 0.40 | 5.8±  2.7 | 6.1±  3.8 | 0.56 | | 5.5±  3.2 | 6.3±  3.5 | 0.40 | 5.9±  3.7 | 5.8±  2.6 | 0.76 |
| CSF Cer C20:0, nM  (N = 91) | 0.6±  0.3 | 0.8±  0.5 | 0.14 | 0.7±  0.4 | 0.7±  0.4 | 0.79 | 0.7±  0.4 | 0.7±  0.4 | 0.93 | | 0.7±  0.4 | 0.7±  0.4 | 0.79 | 0.7±  0.5 | 0.7±  0.3 | 0.79 |
| CSF Cer C22:0, nM  (N = 91) | 0.9±  0.7 | 1.3±  1.2 | 0.095 | 1.2±  1.1 | 0.8±  0.6 | 0.43 | 1.1±  0.9 | 1.1±  1.1 | 0.45 | | 1.0±  1.0 | 1.2±  1.1 | 0.66 | 1.2±  1.1 | 1.0±  0.8 | 0.45 |
| CSF Cer C24:1, nM  (N = 91) | 2.9±  2.8 | 4.5±  4.7 | 0.12 | 4.0±  4.4 | 2.8±  2.0 | 0.30 | 3.5±  3.3 | 4.0±  4.6 | 0.30 | | 3.6±  4.2 | 3.9±  4.0 | 0.71 | 4.1±  4.5 | 3.1±  3.1 | 0.30 |
| CSF Cer C24:0, nM  (N = 91) | 1.9±  1.2 | 1.8±  1.2 | 0.93 | 1.9±  1.3 | 1.6±  0.8 | 0.93 | 1.9±  1.4 | 1.8±  1.1 | 0.93 | | 2.1±  1.3 | 1.7±  1.1 | 0.43 | 1.9±  1.1 | 1.9±  1.5 | 0.93 |
| CSF GlcCer C18:0, nM  (N = 44) | 0.3±  0.2 | 0.4±  0.2 | 0.13 | 0.3±  0.2 | 0.4±  0.2 | 0.13 | 0.3±  0.2 | 0.3±  0.2 | 0.16 | | 0.2±  0.2 | 0.4±  0.2 | 0.13 | 0.3±  0.2 | 0.3±  0.2 | 0.93 |
| CSF GlcCer C22:0, nM  (N = 53) | 8.7±  4.7 | 7.4±  2.9 | 0.55 | 7.8±  3.3 | 8.7±  5.3 | 0.55 | 8.1±  4.2 | 7.9±  3.6 | 0.99 | | 8.5±  4.4 | 7.6±  3.4 | 0.55 | 8.2±  4.1 | 7.5±  3.5 | 0.55 |
| CSF GlcCer C24:1, nM  (N = 53) | 1.8±  1.0 | 1.6±  0.7 | 0.64 | 1.6±  0.6 | 2.0±  1.2 | 0.32 | 1.8±  1.0 | 1.6±  0.6 | 0.76 | | 1.8±  0.9 | 1.6±  0.7 | 0.32 | 1.8±  1.0 | 1.5±  0.6 | 0.32 |
| CSF GlcCer C24:0, nM  (N = 53) | 13.7±  7.6 | 12.0±  4.2 | 0.60 | 12.3±  5.7 | 14.4±  6.9 | 0.60 | 12.7±  6.0 | 12.9±  6.0 | 0.70 | | 14.0±  6.6 | 11.8±  5.4 | 0.17 | 13.0±  6.7 | 12.6±  5.3 | 0.70 |
| **AMP-PD (N = 2,172)** | | | | | | | | | | | | | | | | |
| GCase, %  (N = 49) | 6.4±  2.5 | 5.9±  2.6 | 0.45 | 6.3±  2.6 | 5.0±  1.3 | 0.44 | 5.6±  2.3 | 6.4±  2.6 | 0.45 | | NA | | | 6.5±  2.4 | 5.9±  2.6 | 0.51 |
| Plasma Total Cer, ug/ml  (N = 175) | 4.7±  1.4 | 4.7±  1.6 | 0.81 | 4.8±  1.6 | 4.6±  1.2 | 0.81 | 4.5±  1.3 | 4.9±  1.7 | 0.22 | | NA | | | 4.7±  1.4 | 4.7±  1.6 | 0.81 |
| Plasma Total GlcCer, ug/ml  (N = 212) | 4.8±  1.7 | 4.7±  1.7 | 0.82 | 4.7±  1.7 | 4.8±  1.8 | 0.82 | 4.7±  1.7 | 4.7±  1.7 | 0.82 | | NA | | | 4.7±  1.6 | 4.7±  1.8 | 0.82 |
| Plasma Total LacCer, ug/ml   (N = 175) | 4.5±  1.4 | 4.3±  1.7 | 0.67 | 4.4±  1.4 | 4.2±  2.1 | 0.67 | 4.2±  1.4 | 4.6±  1.7 | 0.32 | | NA | | | 4.4±  1.4 | 4.4±  1.8 | 0.68 |
| Plasma Total SM, ug/ml   (N = 175) | 221.1±  49.2 | 204.3±  44.9 | 0.023* | 212.8±  47.4 | 204.0±  46.5 | 0.53 | 206.3±  45.8 | 215.7±  48.4 | 0.16 | | NA | | | 209.3±  46.8 | 213.8±  48.1 | 0.78 |

Data are presented as mean ± standard deviation; TUEPAC-DESCRIBE-PD, P value corrected by sex, age at visit, disease duration and *GBA1* mutation status; AMP-PD, P value corrected by sex, age at visit, and *GBA1* mutation status; Risk indicates PD patients carrying the risk allele; Pro indicates PD patients carrying non-risk allele. P < 0.05 is considered statistically significant (*). P values were corrected for multiple testing across five genetic variants using the Benjamini–Hochberg method to control the false discovery rate.

Abbreviation: GCase, Glucocerebrosidase; CSF, cerebrospinal fluid; Sph, sphingosine; SPA, sphinganine; S1P, sphingosine-1-phosphate; Cer, ceramides; GlcCer, glucosylceramides; LacCer, lactosylceramides; SM, sphingomyelin.

Table S5. Correlations between Clinical outcomes and sphingolipids profiles in TUEPAC-DESCRIBE-PD.

|  | UPDRS-III (N = 2,046) | | MoCA (N = 2,092) | | BDI-II (N = 1,837) | |
| --- | --- | --- | --- | --- | --- | --- |
|  | rho | FDR corrected *P* | rho | FDR corrected *P* | rho | FDR corrected *P* |
| GCase, % (N = 124) | -0.18 | 0.17 | 0.12 | 0.45 | -0.12 | 0.45 |
| CSF_Sph_nM (N = 90) | 0.075 | 0.77 | -0.18 | 0.25 | 0.062 | 0.80 |
| CSF_SPA_nM (N = 73) | -0.15 | 0.46 | 0.071 | 0.79 | -0.14 | 0.52 |
| CSF_S1P_nM (N = 42) | -0.0030 | 0.99 | 0.15 | 0.62 | -0.36 | 0.078 |
| CSF_C14_cer_nM (N = 91) | 0.26 | 0.058 | -0.081 | 0.75 | -0.013 | 0.96 |
| CSF_C16_cer_nM (N = 91) | -0.062 | 0.80 | 0.0057 | 0.98 | -0.025 | 0.93 |
| CSF_C18_cer_nM (N = 91) | -0.040 | 0.86 | 0.0070 | 0.98 | -0.0072 | 0.98 |
| CSF_C20_cer_nM (N = 91) | 0.024 | 0.93 | -0.029 | 0.91 | -0.0034 | 0.98 |
| CSF_C22_cer_nM (N = 91) | 0.073 | 0.77 | -0.13 | 0.50 | 0.052 | 0.81 |
| CSF_C24_1_cer_nM (N = 91) | 0.0041 | 0.98 | -0.080 | 0.75 | 0.023 | 0.94 |
| CSF_C24_cer_nM (N = 91) | 0.034 | 0.89 | -0.21 | 0.17 | 0.075 | 0.77 |
| CSF_Glc_C18_Cer_nM (N = 44) | -0.16 | 0.61 | 0.19 | 0.49 | -0.13 | 0.75 |
| CSF_Glc_C22_Cer_nM (N = 53) | -0.0062 | 0.98 | -0.12 | 0.71 | 0.081 | 0.80 |
| CSF_Glc_C24_1_Cer_nM (N = 53) | -0.19 | 0.45 | -0.069 | 0.81 | -0.012 | 0.98 |
| CSF_Glc_C24_Cer_nM (N = 53) | -0.088 | 0.79 | -0.10 | 0.76 | 0.14 | 0.62 |

FDR corrected *P*, P values were corrected for multiple testing across all correlation tests using the Benjamini–Hochberg method to control the false discovery rate.

Table S6. Correlations between lysosomal protein and clinical outcomes, sphingolipids profiles in TUEPAC-DESCRIBE-PD.

|  | LAMP2 (N = 357) | | LIMP2 (N = 135) | | CTSB (N = 135) | |
| --- | --- | --- | --- | --- | --- | --- |
|  | rho | FDR corrected *P* | rho | FDR corrected *P* | rho | FDR corrected *P* |
| LAMP2 (N = 357) |  |  |  |  |  |  |
| LIMP2 (N = 135) | 0.14 | 0.70 |  |  |  |  |
| CTSB (N = 135) | -0.21 | 0.45 | -0.27 | 9.83e-03* |  |  |
| MoCA (N = 2,092) | 0.054 | 0.60 | -0.12 | 0.46 | 0.11 | 0.46 |
| BDI-II (N = 1,837) | 0.039 | 0.77 | 0.041 | 0.89 | -0.21 | 0.35 |
| UPDRS-III (N = 2,046) | -0.14 | 0.034* | 0.24 | 0.024* | -0.15 | 0.22 |
| GCase, % (N = 124) | 0.16 | 0.45 | -0.078 | 0.81 | 0.086 | 0.80 |
| CSF_Sph_nM (N = 90) | 0.015 | 0.96 | -0.11 | 0.78 | 0.12 | 0.77 |
| CSF_SPA_nM (N = 73) | 0.080 | 0.79 | -0.058 | 0.90 | -0.0023 | 0.99 |
| CSF_S1P_nM (N = 42) | 0.077 | 0.83 | -0.24 | 0.59 | 0.26 | 0.52 |
| CSF_C14_cer_nM (N = 91) | -0.048 | 0.84 | -0.072 | 0.84 | 0.17 | 0.62 |
| CSF_C16_cer_nM (N = 91) | -0.012 | 0.98 | 0.070 | 0.84 | -0.024 | 0.96 |
| CSF_C18_cer_nM (N = 91) | 0.14 | 0.50 | 0.43 | 0.044* | -0.086 | 0.81 |
| CSF_C20_cer_nM (N = 91) | -0.10 | 0.67 | 0.13 | 0.75 | -0.032 | 0.95 |
| CSF_C22_cer_nM (N = 91) | -0.29 | 0.044* | -0.030 | 0.95 | 0.10 | 0.80 |
| CSF_C24_1_cer_nM (N = 91) | -0.39 | 2.85e-03* | 0.090 | 0.81 | 0.084 | 0.81 |
| CSF_C24_cer_nM (N = 91) | -0.43 | 5.87e-04* | -0.077 | 0.83 | 0.032 | 0.95 |
| CSF_Glc_C18_Cer_nM (N = 44) | -0.14 | 0.70 | 0.060 | 0.95 | -0.22 | 0.77 |
| CSF_Glc_C22_Cer_nM (N = 53) | -0.52 | 1.25e-03* | -0.095 | 0.85 | 0.12 | 0.81 |
| CSF_Glc_C24_1_Cer_nM (N = 53) | -0.52 | 1.52e-03* | 0.15 | 0.79 | -0.12 | 0.81 |
| CSF_Glc_C24_Cer_nM (N = 53) | -0.44 | 0.010* | -0.075 | 0.89 | -0.25 | 0.59 |

FDR corrected *P*, P values were corrected for multiple testing across all correlation tests using the Benjamini–Hochberg method to control the false discovery rate.

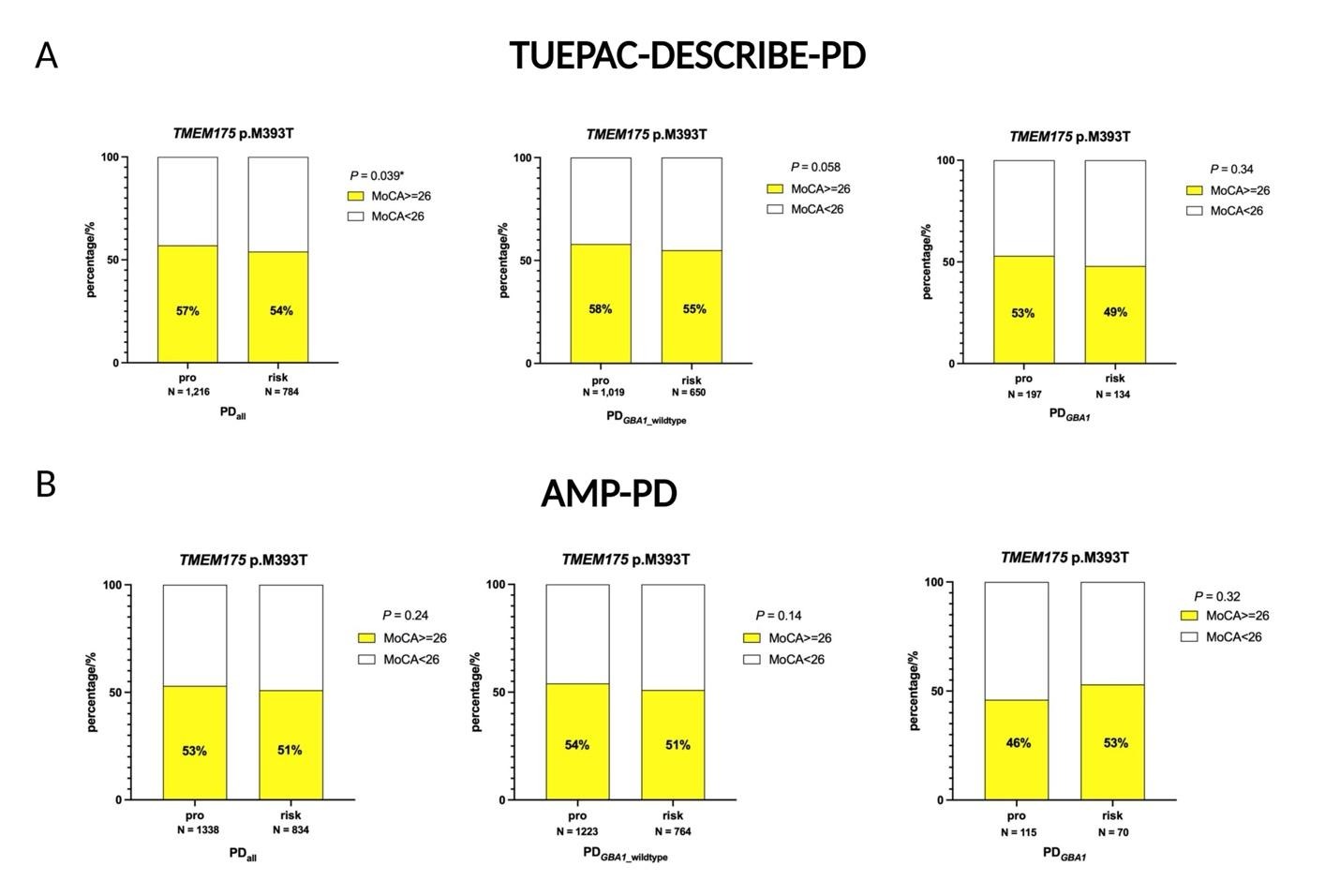

Figure S1. Associations between *TMEM175* p.M393T and cognitive function (MoCA scale) using logistic regression analysis. A) TUEPAC-DESCRIBE-PD: Proportion of individuals with MoCA scores ≥ 26 between protective (pro) and risk allele carriers of the *TMEM175* p.M393T variant across PD_all_, PD*_GBA1_*__wildtype_ and PD*_GBA1_*; B) AMP-PD: Proportion of individuals with MoCA scores ≥ 26 between protective (pro) and risk allele carriers of the *TMEM175* p.M393T variant, stratified by PD_all_, PD*_GBA1_*__wildtype_ and PD*_GBA1_*.

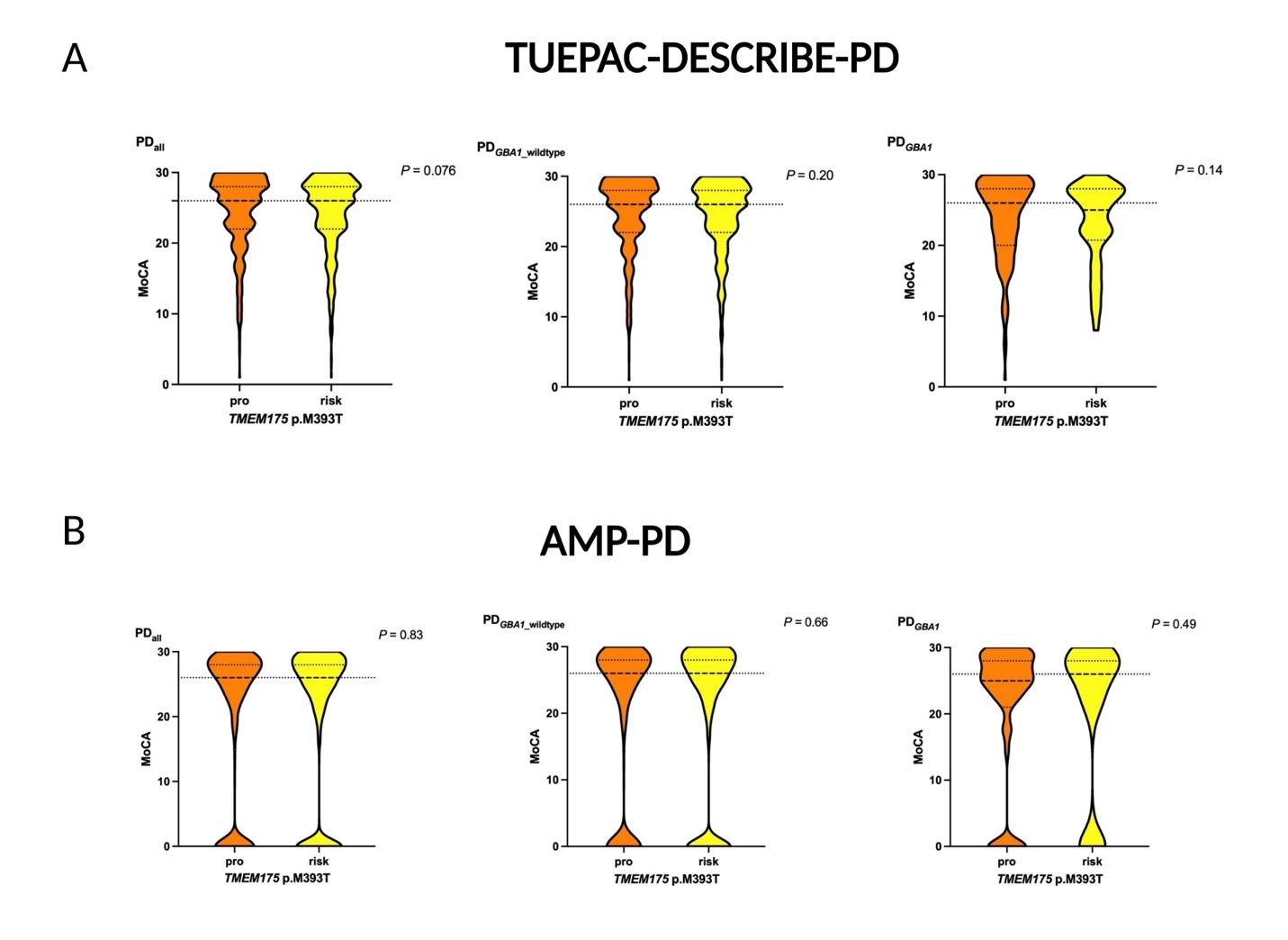

Figure S2. Associations between *TMEM175* p.M393T and cognitive function (MoCA scale) using linear regression analysis. A) TUEPAC-DESCRIBE-PD: MoCA scores distribution between protective (pro) and risk allele carriers of the *TMEM175* p.M393T variant across PD_all_, PD*_GBA1_*__wildtype_ and PD*_GBA1_*; B) AMP-PD: MoCA scores distribution between protective (pro) and risk allele carriers of the *TMEM175* p.M393T variant, stratified by PD_all_, PD*_GBA1_*__wildtype_ and PD*_GBA1_*.
